# Pre-fracture Anemia Is Associated with Nonunion Following Tibia or Femur Fractures: A Retrospective Cohort Study

**DOI:** 10.64898/2026.07.16.26358267

**Authors:** Christophe Merceron, Sanidhya Singh, Daniel G. Whitney, Andrea I. Alford, Shikha Sachdeva, Rami Khoriaty, Brandi Hartley, Annemarie Lang

## Abstract

Fracture nonunion remains a major cause of morbidity, yet patient-specific factors associated with impaired healing remain incompletely characterized. Anemia has been associated with adverse orthopaedic outcomes, but its relationship with fracture nonunion is poorly understood. We examined whether pre-fracture anemia, anemia burden, and clinically relevant anemia subtypes were associated with nonunion following tibial or femoral fractures. Using commercial and Medicare fee-for-service claims from 2016 through 2023, we identified adults aged 19 years or older with a tibial or femoral fracture, continuous enrollment during the preceding year and for at least six months after fracture, and no baseline cancer. Pre-fracture anemia was evaluated as any anemia, the number of distinct anemia diagnoses, and nutritional, hemolytic, aplastic, and other anemia subgroups. Nonunion occurring six to eighteen months after fracture was assessed using incidence rates and multivariable-adjusted hazard models. Among 326,673 adults, 149,704 had pre-fracture anemia and 176,969 did not. The crude incidence of nonunion was 42% higher among individuals with anemia than among those without anemia (incidence rate ratio, 1.42; 95% confidence interval, 1.32 to 1.53) and increased with greater anemia burden. After adjustment for demographic and clinical characteristics, including prior fractures at other anatomical sites, pre-fracture anemia remained associated with nonunion following tibial and femoral fractures, with hazard ratios of 1.83 (95% confidence interval, 1.54 to 2.18) and 1.38 (95% confidence interval, 1.26 to 1.50), respectively. Associations were also observed for nutritional and other anemias, whereas estimates for hemolytic and aplastic anemias were limited by few nonunion events. Within the femur, the association was strongest for distal fractures. These findings demonstrate that pre-fracture anemia is independently associated with nonunion. The increase in risk with greater anemia burden and findings across evaluable subgroups suggest that pre-fracture anemia may help identify patients at increased risk of impaired fracture healing.

## Introduction

Bone fracture healing is generally predictable, yet a substantial proportion of patients experience impaired repair, leading to delayed union or nonunion. With an estimated 178 million fractures occurring worldwide each year and 10‒15% failing to heal appropriately^(1)^, fracture nonunion remains a significant source of morbidity and healthcare burden. Despite advances in surgical and biologic therapies, current strategies remain largely reactive, focusing on treating established healing failure rather than preventing it. Although numerous risk factors have been identified, they remain incompletely characterized, and robust predictive markers are limited. There is a critical unmet need for mechanistically informed approaches that identify patients at risk and enable early, targeted intervention.

Anemia represents a highly prevalent yet underappreciated systemic risk factor for impaired musculoskeletal regeneration. Affecting approximately 24% of the global population (1.9 billion people)^(2)^, with iron-deficiency anemia accounting for nearly two-thirds of cases, anemia disproportionately impacts populations already at elevated fracture risk^(3)^. In the United States, anemia is more common among women, older adults, and individuals with chronic disease, groups that are also at increased risk for fracture nonunion^(1,3)^. Despite its prevalence and treatability, anemia is not routinely incorporated into fracture risk assessment, largely because its mechanistic contribution to bone repair remains poorly defined.

Anemia subtypes, such as nutritional, hemolytic, and aplastic anemia, reflect diverse underlying pathophysiologic processes that may differentially influence fracture healing. Clinical studies increasingly associate anemia with adverse orthopaedic outcomes: preoperative anemia predicts higher rates of postoperative complications, transfusion requirements, non-home discharge, and mortality following fracture fixation and joint reconstruction^(4,5)^. Similarly, injury-associated anemia is nearly universal following operative fracture, with functional iron deficiency present in the majority of patients yet frequently underdocumented^(6)^; when persistent, post-injury anemia is associated with prolonged hospitalization, increased complications, and worse recovery trajectories^(7–9)^. Iron indices in these patients frequently suggest functional iron deficiency, indicating iron sequestration rather than absolute depletion^(6)^. Iron-deficiency anemia has also been implicated as a contributor to osteoporosis, even prior to menopause^(10)^. Other disorders of erythropoiesis, including sickle cell disease and myelodysplastic syndromes, are associated with skeletal abnormalities, impaired bone integrity, and increased fracture risk^(11–14)^. However, the mechanisms by which anemia influences fracture healing remain poorly understood.

Most existing studies focus on bone mineral density or structural bone parameters, with limited investigation into anemia as a predictor of fracture healing outcomes. To address this gap, we examined whether pre-fracture anemia is associated with an increased risk of nonunion following tibial or femoral fractures. We hypothesized that patients with anemia prior to fracture would have a significantly higher incidence of nonunion. This retrospective cohort study utilized commercial and Medicare fee-for-service claims data from 2016 to 2023, including adults ≥19 years old with tibia or femur fractures and continuous enrollment before and after injury. Pre-fracture anemia within one year was evaluated as the primary exposure, and associations with nonunion (6‒18 months post-fracture) were assessed using incidence rates and adjusted hazard models.

## Methods

### Data source

This was a retrospective cohort study that accessed administrative claims data for research purposes. Claims from January 1, 2016, through December 31, 2023, were ascertained from two separate databases: the Commercial Database from the IBM® Watson Health MarketScan® Research Databases and a 20% random sample of the Medicare fee-for-service database (Part A and B) from the Centers for Medicare & Medicaid Services. These databases were harmonized in this study as the Commercial database is representative of the general working non-elderly population <65 years of age while the Medicare database is representative of the elderly population ≥65 years of age. Thus, by harmonizing datasets, this study developed a large, heterogenous cohort across the adult life course with geographic diversity, enhancing representation and generalizability of findings.

The IBM® MarketScan® Commercial Database includes health insurance related data from the private-sector encompassing ∼350 payers across the U.S. and represents individuals that are insured employees, continuants of the Consolidated Omnibus Budget Reconciliation Act, and non-Medicare eligible dependents that have insurance coverage based on employer-sponsored plans. Medicare is a federal program providing health insurance to the elderly, as well as elderly and non-elderly with specific disabilities or with end-stage renal disease. Medicare beneficiaries can have dual eligibility with Medicaid, but for those with dual eligibility, Medicare pays first for services that are covered by Medicare and Medicaid, including the services needed to conduct this study. The Institutional Review Board of the University of Michigan waived ethical approval for this work. The databases were de-identified prior to being administered to researchers, and patient consent was not required.

### Cohort

See **Supplementary Figure 1** for the flow chart of inclusion/exclusion criteria. Individuals were included in this study if they experienced an incident fracture at the tibia or femur between Jan. 1, 2017 (to allow for a 1-year baseline period) and June 30, 2022 (to allow up to 18-months of follow-up) with at least 1- year of a “clean” period, i.e., no evidence of a tibia or femur fracture in the year preceding. Additional inclusion criteria were 19 years of age or older by the date of the qualifying fracture, continuous health plan enrollment for at least 1 year prior to the qualifying fracture date, and at least 6 months of continuous health plan enrollment after the qualifying fracture date. This study forced post-fracture enrollment for at least 6 months as this was the start of the at-risk window for the outcome, from 6-months post-fracture through 18-months post-fracture. Exclusion criteria were evidence of any type of cancer in the 1-year prior to the fracture date, evidence of the outcome up to 6-months post-fracture and having a fracture at both the tibia and femur for the same fracture event as this may indicate polytrauma.

### Exposure

Of the included cohort with a qualifying tibia or femur fracture, baseline evidence of anemia was defined as having at least 1 claim with a pertinent International Classification of Diseases, Tenth Revision (ICD- 10) code for any type of anemia including 14 individual conditions (ICD-10 codes D50.x-D53.x and D55.x- D64.x) during the 1-year baseline period. Evidence of anemia on or following the qualifying fracture date was not considered in this study as the fracture event can lead to new-onset anemia, potentially masking associations of interest. Anemia was examined in three ways to improve the understanding of the effect of anemia on the outcome. First, anemia was examined as presents or not present to any type of anemia. Second, anemia burden was evaluated by counting the number of distinct anemia conditions documented during the one-year baseline period to determine whether a greater number of anemia diagnoses was associated with progressively higher nonunion risk. Because relatively few individuals had large numbers of anemia diagnoses, this variable was categorized as 0, 1, 2, or 3 or more distinct anemia conditions. Third, anemia was examined by the broad grouping that clinically differentiates the various types to assess for variation in effects by different anemia groups, including nutritional anemias (ICD-10 codes D50.x-D53.x), hemolytic anemias (ICD-10 codes D55.x-D59.x), aplastic anemias (ICD-10 codes D60.x- D61.x) and other anemias (ICD-10 codes D62.x-D64.x).

### Outcomes

The outcome in this study was nonunion at the same fracture site, i.e., nonunion of the tibia or nonunion of the femur. The first claim with a pertinent ICD-10 code for site-specific nonunion was considered the qualifying outcome event. Individuals were excluded from the analysis if they had evidence of the outcome within 6 months of the qualifying fracture date. Six months were selected for clinical relevance, as the FDA’s most widely accepted definition of nonunion is a fracture that persists for at least 9 months without signs of healing for 3 months^(15)^, and to avoid the inclusion of false positives or nonunions diagnosed for reasons independent of failed biological healing.

### Descriptive characteristics

Year of the qualifying fracture date, age at the qualifying fracture date, sex, and U.S. region of residence were retrieved from both databases. Insurance type was examined as Commercial, Medicare only, or Medicare + Medicaid dual eligibility. For the Medicare database, the original reason for Medicare entitlement was also retrieved. The Elixhauser Comorbidity Index, a proxy of overall health status, was estimated using the full 1-year baseline period, which is the sum of 30 comorbidities based on diagnostic evidence (1 point per comorbidity) or lack thereof (0 points per comorbidity), allowing a total score range from 0 to 30^(16)^. For this study, blood loss anemia and any kind of deficiency anemia were excluded from the comorbidity index as these ICD-10 codes were included in the anemia exposure, thus overcounting the index score for some in the anemia cohort; the index revised for this study therefore ranged from 0 to 28. Evidence of iron deficiency during the 1-year baseline period was ascertained based on the ICD- 10 code E61.1. The type of fracture was also determined based on the ICD-10 code of the qualifying fracture event, including closed, open, or pathological (including atypical femur fracture) fracture. Baseline evidence of fracture at any site (other than femur and tibia given the inclusion criteria) during the 1-year prior the fracture event was identified using ICD-10 codes. The Commercial database does not provide information about race or ethnicity, and as such, these variables were not assessed in this study.

### Statistical analysis – primary objective

Baseline descriptive characteristics were summarized for the cohorts with vs. without anemia. The crude incidence rate (IR) with 95% confidence intervals (CI) per 1,000 person-years of the outcome was estimated for the cohort with and without anemia and by the anemia sub-groups (i.e., count of co-occurring anemic conditions, clinically distinct types of anemia). The crude IR ratio (IRR) was estimated by comparing the IR of the anemia and anemia sub-groups to the IR of the cohort without anemia. The cumulative incidence of the outcomes using the Fine-Gray approach were plotted to visualize the time-varying pattern^(17)^. Individuals were followed until the date of the outcome event, lost to follow-up, or end of the 18-month follow-up period, whichever came first.

Cox proportional hazards regression was used to estimate the hazard ratio (HR) of the outcome comparing the anemia and anemia sub-groups to the cohort without anemia. To first determine if the tibia and femur fracture cohorts could be combined or if effects needed to be stratified by fracture site, pre-modelling tested for effect modification between each cohort variable and fracture site. There was strong evidence of effect modification by fracture sites regardless of how anemia was examined (i.e., present/not present, count of co-occurring anemic conditions, clinically distinct types of anemia) (*P* for interaction, ranged from <0.001 to 0.009). Therefore, sub-group estimates were generated by fracture site in all models before and after adjusting for age (as continuous), sex, U.S. region of residence, insurance type, Elixhauser Comorbidity Index score, iron deficiency, fracture type of the qualifying fracture event, and baseline fracture at any site. Effect modification by each covariate was tested in separate models using a 3-way interaction (including main effects) between the cohort variable, fracture site, and the covariate being tested (e.g., age, sex). The proportional hazards assumption was tested based on the weighted Schoenfeld residuals.

### Statistical analysis – secondary objective

Bone marrow composition varies across anatomical regions of the tibia and femur, with haematopoietically active red marrow and metabolically inactive yellow marrow distributed differentially across sites in a manner that changes with age, a distinction that may influence the biological environment of fracture healing and susceptibility to nonunion. Therefore, a secondary analysis was conducted using Cox regression to assess the effect of anemia on nonunion by fracture region separately for the tibia and femur, including the proximal, shaft, distal, and unspecified sites. For these analyses, effect modification between each cohort variable and fracture region was tested first. If there was no evidence of effect modification (*P*>0.10), the fracture region was set as the primary exposure, and the HR was estimated before and after adjusting for the same covariates noted above in the primary objective in addition to anemia (as present/not present). If there was evidence of effect modification (*P*<0.10), then sub-group estimates were generated by fracture region before and after adjusting for the covariates noted above in the primary objective.

As part of the Data Use Agreement, estimates with n=1 to n=10 cases were suppressed for patient de-identification purposes. Analyses were performed using SAS version 9.4 (Cary, NC) and P<0.05 (two-tailed) was considered statistically significant.

## Results

### Descriptive characteristics

Baseline descriptive characteristics of the cohort with anemia (n=149,704) and without anemia (n=176,969) is presented in **Table 1**. In general, the cohort with anemia was older on average by 9.0 years and had a slightly higher proportion of women (73.5% vs. 65.8%), while the distribution by U.S. region of residence was relatively similar between groups. The anemia cohort was also sicker overall, as evidenced by a higher Elixhauser Comorbidity Index score (median of 3 vs. 1), and was less likely to be covered by commercial insurance (4.9% vs. 24.8%). Additionally, those with anemia were more likely to have pre-existing iron deficiency (1.6% vs. 0.3%) and more frequently sustained a fracture at the femur (88.4% vs. 67.1%) rather than the tibia (11.6% vs. 32.9%).

**Table 1.**
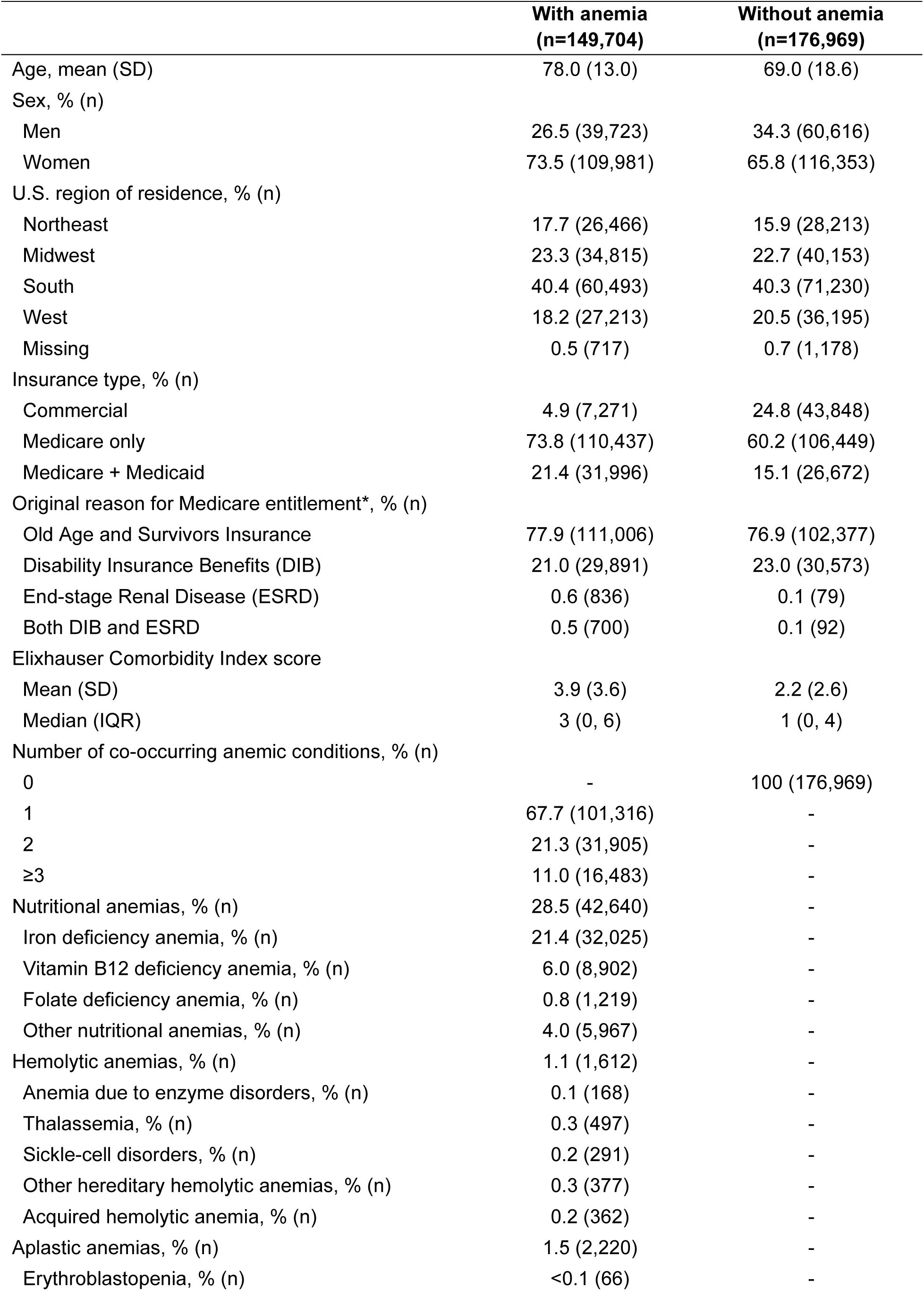
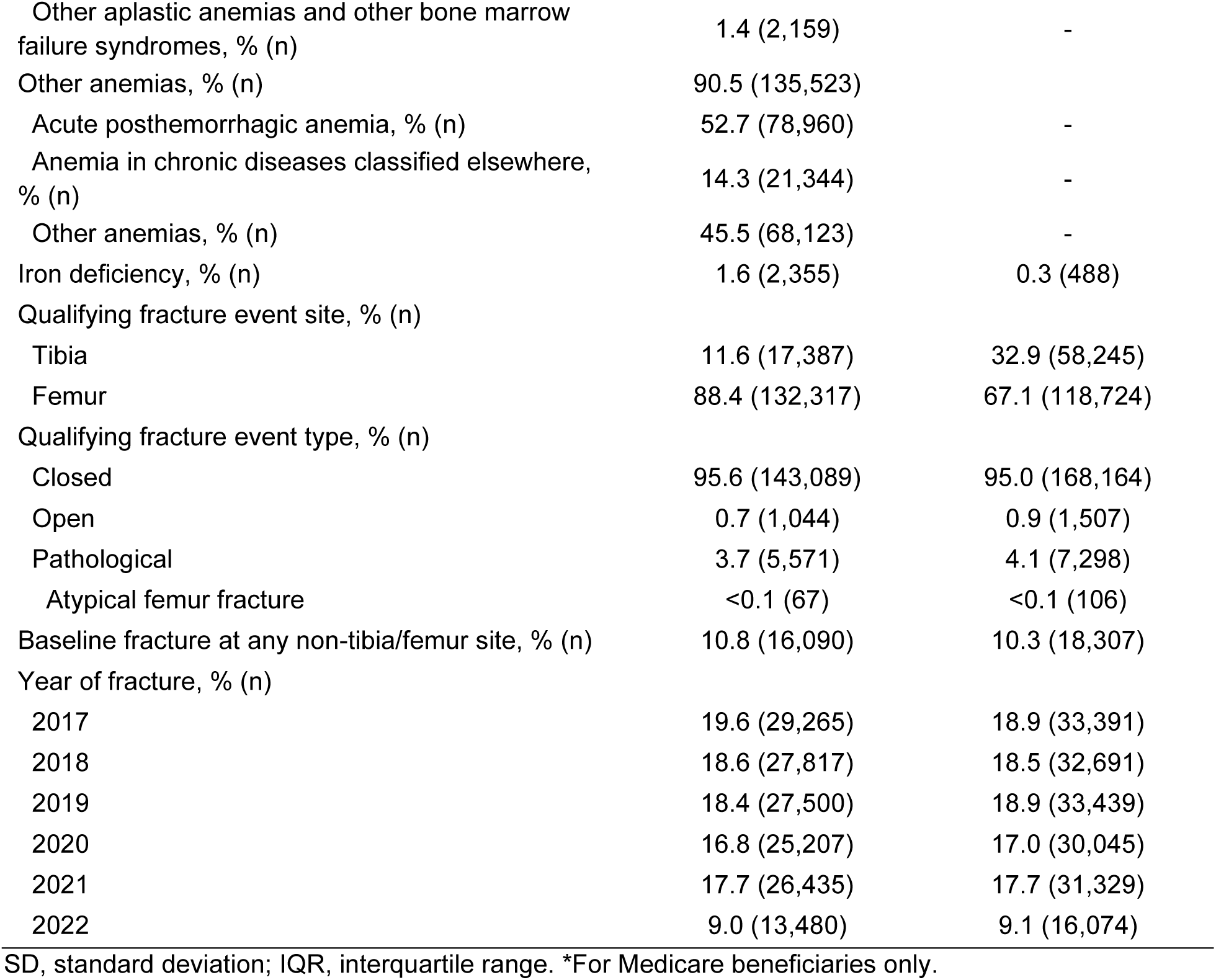
Baseline descriptive characteristics of the cohorts with and without anemia.

Among the cohort with anemia, nearly 2/3rds had only 1 of the 14 studied anemic conditions (67.7%) while 21.3% had 2 co-occurring anemic conditions and 11.0% had 3 or more co-occurring anemic conditions. Of the clinically distinct types of anemia, the most common grouping was other anemias (90.5% of the cohort with anemia) followed by nutritional anemias (28.5%), aplastic anemias (1.5%), and hemolytic anemias (1.1%).

### Primary objective

The censor reason, IR, and IRR of nonunion for the combined and site-specific fracture sites are presented in **Table 2** and the cumulative incidence plot is presented in **Figure 1**. For the combined fracture sites (tibia or femur), the IR (95% CI) per 1,000 person-years of nonunion was 7.9 (7.5-8.3) for the cohort with anemia and 5.6 (5.3-5.9) for the cohort without anemia; the IRR was 42% higher (IRR=1.42; 95% CI=1.32-1.53). The crude IR and IRR were higher for every additional co-occurring anemic condition and was slightly higher for the nutritional anemias (IR=8.6, IRR vs. no anemia = 1.54) and aplastic anemias (IR=9.9, IRR vs. no anemia = 1.78) followed by hemolytic anemias (IR=8.4, IRR vs. no anemia = 1.51) and other anemias (IR=7.3, IRR vs. no anemia = 1.32). The findings of an elevated crude incidence of nonunion among the anemia and anemia sub-groups were similar after stratifying by fracture site, except that the tibia fracture site had a greater disparity than the femur fracture site.

**Figure 1:**
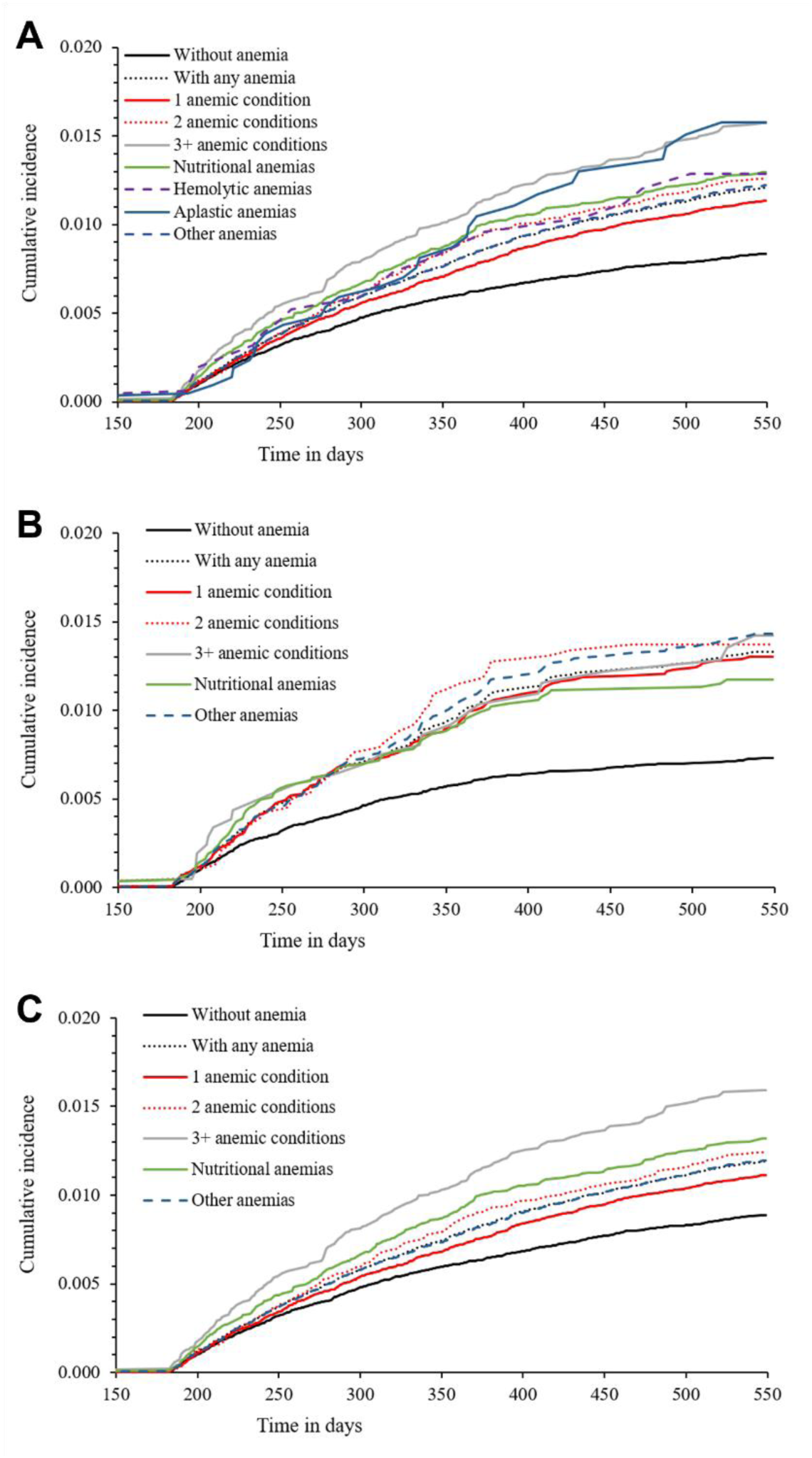
Cumulative incidence plot of nonunion up to 18-months following a (A) combined tibia or femur fracture, (B) tibia fracture only, or (C) femur fracture only for the cohorts with and without anemia and by anemia sub-groups. The start of the at-risk window began at 6-months post-fracture. To enhance visualization, the x-axis starts at 150 days. Data are omitted from the hemolytic and aplastic anemia cohorts for patient de-identification purposes in B and C because at least one site had N<11 events.

**Table 2.** Censor reason, incidence rate (IR), and IR ratio (IRR) of nonunion up to 18-months following a tibia or femur fracture for the cohorts with vs. without anemia and by anemia sub-groups.

|  | Censor reason |  |  |  | Person-time | Crude IR | Crude IRR |
| --- | --- | --- | --- | --- | --- | --- | --- |
|  | Sample size | End of follow-up | Lost to follow-up | Event |  |  |  |
|  | N | % (n) | % (n) | % (n) | Person-years | IR (95% CI) | IRR (95% CI) |
| **Fracture at the tibia or femur** |  |  |  |  |  |  |  |
| Without anemia | 176,969 | 72.0 (127,439) | 27.2 (48,198) | 0.8 (1,332) | 239,538 | 5.6 (5.3, 5.9) | Reference |
| With any anemia | 149,704 | 67.3 (100,713) | 31.7 (47,421) | 1.1 (1,570) | 198,461 | 7.9 (7.5, 8.3) | **1.42 (1.32, 1.53)** |
| Number of co-occurring anemic conditions |  |  |  |  |  |  |  |
| 1 | 101,316 | 68.6 (69,461) | 30.5 (30,851) | 1.0 (1,004) | 135,134 | 7.4 (7.0, 7.9) | **1.34 (1.23, 1.45)** |
| 2 | 31,905 | 65.7 (20,967) | 33.2 (10,592) | 1.1 (346) | 41,962 | 8.2 (7.4, 9.1) | **1.48 (1.32, 1.67)** |
| ≥3 | 16,483 | 62.4 (10,285) | 36.3 (5,978) | 1.3 (220) | 21,365 | 10.3 (8.9, 11.7) | **1.85 (1.61, 2.14)** |
| Nutritional anemias | 42,640 | 65.6 (27,970) | 33.3 (14,190) | 1.1 (480) | 56,095 | 8.6 (7.8, 9.3) | **1.54 (1.39, 1.71)** |
| Hemolytic anemias | 1,612 | 69.2 (1,115) | 29.7 (479) | 1.1 (18) | 2,143 | 8.4 (4.5, 12.3) | 1.51 (0.95, 2.40) |
| Aplastic anemias | 2,220 | 59.7 (1,325) | 39.1 (867) | 1.3 (28) | 2,831 | 9.9 (6.2, 13.6) | **1.78 (1.22, 2.59)** |
| Other anemias | 135,523 | 67.2 (91,020) | 31.8 (43,069) | 1.1 (1,434) | 195,629 | 7.3 (7, 7.7) | **1.32 (1.22, 1.42)** |
| **Fracture at the tibia** |  |  |  |  |  |  |  |
| Without anemia | 58,245 | 75.4 (43,937) | 23.9 (13,914) | 0.7 (394) | 79,827 | 4.9 (4.4, 5.4) | Reference |
| With any anemia | 17,387 | 70.8 (12,314) | 28.0 (4,866) | 1.2 (207) | 23,353 | 8.9 (7.7, 10.1) | **1.80 (1.52, 2.12)** |
| Number of co-occurring anemic conditions |  |  |  |  |  |  |  |
| 1 | 11,379 | 71.9 (8,176) | 27.0 (3,070) | 1.2 (133) | 15,352 | 8.7 (7.2, 10.1) | **1.76 (1.44, 2.14)** |
| 2 | 3,891 | 69.8 (2,717) | 28.9 (1,126) | 1.2 (48) | 5,206 | 9.2 (6.6, 11.8) | **1.87 (1.38, 2.52)** |
| ≥3 | 2,117 | 67.1 (1,421) | 31.7 (670) | 1.2 (26) | 2,795 | 9.3 (5.7, 12.9) | **1.88 (1.27, 2.80)** |
| Nutritional anemias | 6,850 | 71.2 (4,877) | 27.7 (1,900) | 1.1 (73) | 9,217 | 7.9 (6.1, 9.7) | **1.60 (1.25, 2.06)** |
| Hemolytic anemias | 339 | 74.9 (254) | * | * | * | * | 1.33 (0.43, 4.14) |
| Aplastic anemias | 334 | 66.5 (222) | * | * | * | * | 1.83 (0.68, 4.91) |
| Other anemias | 14,485 | 70.1 (10,155) | 28.6 (4,146) | 1.3 (184) | 19,393 | 9.5 (8.1, 10.9) | **1.92 (1.61, 2.29)** |
| **Fracture at the femur** |  |  |  |  |  |  |  |
| Without anemia | 118,724 | 70.3 (83,502) | 28.9 (34,284) | 0.8 (938) | 159,711 | 5.9 (5.5, 6.2) | Reference |
| With any anemia | 132,317 | 66.8 (88,399) | 32.2 (42,555) | 1.0 (1,363) | 175,107 | 7.8 (7.4, 8.2) | **1.33 (1.22, 1.44)** |
| Number of co-occurring anemic conditions |  |  |  |  |  |  |  |
| 1 | 89,937 | 68.1 (61,285) | 30.9 (27,781) | 1.0 (871) | 119,781 | 7.3 (6.8, 7.8) | **1.24 (1.13, 1.36)** |
| 2 | 28,014 | 65.2 (18,250) | 33.8 (9,466) | 1.1 (298) | 36,756 | 8.1 (7.2, 9.0) | **1.38 (1.21, 1.57)** |
| ≥3 | 14,366 | 61.7 (8,864) | 37.0 (5,308) | 1.4 (194) | 18,570 | 10.4 (9.0, 11.9) | **1.78 (1.52, 2.08)** |
| Nutritional anemias | 35,790 | 64.5 (23,093) | 34.3 (12,290) | 1.1 (407) | 46,878 | 8.7 (7.8, 9.5) | **1.48 (1.32, 1.66)** |
| Hemolytic anemias | 1,273 | 67.6 (861) | * | * | * | * | 1.51 (0.91, 2.52) |
| Aplastic anemias | 1,886 | 58.5 (1,103) | * | <1.5%** | * | * | **1.71 (1.14, 2.56)** |
| Other anemias | 121,038 | 66.8 (80,865) | 32.2 (38,923) | 1.0 (1,250) | 160,182 | 7.8 (7.4, 8.2) | **1.33 (1.22, 1.45)** |

CI, confidence interval. Statistically significant associations at P<0.05 are bolded to enhance visual interpretation.
\*Data are suppressed due to the limited number of outcome events. \*\*Data are suppressed to prevent back calculation of the number of tibia fractures.

The unadjusted and adjusted HR are presented in **Table 3** for the tibia and femur separately. The HR was significantly elevated for all anemia and anemia sub-groups, except for the hemolytic anemias and aplastic anemias sub-groups, when compared to the cohort without anemia before and after adjusting for covariates. Given the limited number of outcome events for the hemolytic anemias sub- group (n=18) and aplastic anemias sub-group (n=28), the adjusted model adjusted for a subset of the covariates including age, sex, and Elixhauser Comorbidity Index score. Notably, the association between anemia and nonunion was stronger among those that sustained a fracture at the tibia than femur. There was no strong evidence that the proportional hazards assumption was violated in any model (all *P*>0.05).

**Table 3.** Hazard ratio (HR) of nonunion up to 18-months following a tibia or femur fracture for the cohorts with vs. without anemia and by anemia sub-groups based on evidence of effect modification by fracture site (tibia, femur).

|  | Tibia |  | Femur |  |
| --- | --- | --- | --- | --- |
|  | Unadjusted<br>HR (95% CI) | Adjusted<br>HR (95% CI) | Unadjusted<br>HR (95% CI) | Adjusted<br>HR (95% CI) |
| Without anemia | Reference | Reference | Reference | Reference |
| With any anemia | **1.80 (1.52, 2.13)** | **1.83 (1.54, 2.18)** | **1.33 (1.22, 1.44)** | **1.38 (1.26, 1.50)** |
| Number of co-occurring anemic conditions |  |  |  |  |
| 1 | **1.76 (1.45, 2.14)** | **1.85 (1.52, 2.27)** | **1.24 (1.13, 1.36)** | **1.36 (1.24, 1.49)** |
| 2 | **1.87 (1.39, 2.53)** | **1.89 (1.39, 2.56)** | **1.39 (1.22, 1.58)** | **1.38 (1.20, 1.58)** |
| ≥3 | **1.90 (1.27, 2.82)** | **1.71 (1.14, 2.57)** | **1.79 (1.53, 2.09)** | **1.51 (1.28, 1.79)** |
| Nutritional anemias | **1.61 (1.25, 2.06)** | **1.61 (1.24, 2.08)** | **1.49 (1.32, 1.67)** | **1.39 (1.23, 1.58)** |
| Hemolytic anemias* | 1.34 (0.43, 4.16) | 1.24 (0.40, 3.86) | 1.52 (0.91, 2.54) | 1.30 (0.78, 2.17) |
| Aplastic anemias* | 1.84 (0.69, 4.93) | 1.67 (0.62, 4.48) | **1.74 (1.16, 2.60)** | 1.38 (0.91, 2.07) |
| Other anemias | **1.93 (1.62, 2.30)** | **1.95 (1.62, 2.34)** | **1.33 (1.22, 1.45)** | **1.38 (1.27, 1.51)** |

CI, confidence interval. The adjusted HR adjusted for age (as continuous), sex, insurance type, U.S. region of residence, Elixhauser Comorbidity Index score, iron deficiency, fracture type of the qualifying fracture event (closed, open, pathological), and baseline fracture at any non tibia/femur site. Statistically significant associations at $P < 0.05$ are bolded to enhance visual interpretation. \*Due to the limited number of outcome events, the adjusted HR adjusted for age (as continuous), sex, and Elixhauser Comorbidity Index score.

There was evidence of a 3-way interaction (cohort x fracture site x covariate) by fracture type for the full anemia cohort (present/not present) and the clinical sub-group nutritional anemias (*P* for interaction, 0.015 and 0.067, respectively) and by the Elixhauser Comorbidity Index score for the clinical sub-group nutritional anemias (*P* for interaction, 0.099). As compared to the cohort without anemia, the adjusted HR was significantly elevated for the cohort with any anemia that sustained a fracture at the femur for all fracture types (closed [HR=1.36], open [HR=2.64], and pathological [HR=1.93]), while the adjusted HR was significantly elevated for the cohort with any anemia that sustained a fracture at the tibia only for closed fractures (HR=1.96), but not for open (HR=1.03) or pathological (HR=0.73) fracture types (**Table 4**). There were similar findings of the fracture type modifier for the nutritional anemias sub- group, but fewer associations reached statistical significance. For the Elixhauser Comorbidity Index score modification, sub-group estimates were generated across the score range by 0, 1, 2, 3, 5, and 7 comorbidities (**Table 4**). Nutritional anemias were associated with a higher rate of nonunion across all scores where the strength of association was slightly stronger for the tibia than femur fracture cohorts, but the HR estimate declined with an increasing score in both tibia and femur fracture cohorts.

**Table 4.** Sub-group estimates based on evidence of effect modification by fracture type and Elixhauser Comorbidity Index score for the outcome, nonunion, up to 18-months following a tibia or femur fracture for the cohorts with vs. without anemia and by anemia sub-groups.

|  | Tibia<br>Adjusted<br>HR (95% CI) | Femur<br>Adjusted<br>HR (95% CI) |
| --- | --- | --- |
| With any anemia (reference: without anemia) |  |  |
| Fracture type |  |  |
| Closed | **1.96 (1.63, 2.36)** | **1.36 (1.24, 1.48)** |
| Open | 1.03 (0.58, 1.83) | **2.64 (1.14, 6.12)** |
| Pathological | 0.73 (0.16, 3.46) | **1.93 (1.10, 3.36)** |
| Nutritional anemias (reference: without anemia) |  |  |
| Fracture type |  |  |
| Closed | **1.77 (1.37, 2.31)** | **1.37 (1.21, 1.55)** |
| Open | 0.49 (0.12, 2.05) | 2.99 (0.90, 9.96) |
| Pathological | * | 2.12 (0.96, 4.65) |
| Nutritional anemias (reference: without anemia) |  |  |
| Elixhauser Comorbidity Index score |  |  |
| 0 comorbidities | **1.71 (1.09, 2.67)** | **1.66 (1.36, 2.02)** |
| 1 comorbidity | **1.65 (1.11, 2.45)** | **1.58 (1.33, 1.88)** |
| 3 comorbidities | **1.54 (1.12, 2.12)** | **1.44 (1.25, 1.65)** |
| 5 comorbidities | **1.44 (1.08, 1.92)** | **1.31 (1.15, 1.49)** |
| 7 comorbidities | 1.34 (0.98, 1.85) | **1.19 (1.02, 1.39)** |

HR, hazard ratio; CI, confidence interval. The adjusted HR adjusted for age (as continuous), sex, insurance type, U.S. region of residence, Elixhauser Comorbidity Index score, iron deficiency, fracture type of the qualifying fracture event (closed, open, pathological), and baseline fracture at any non tibia/femur site. Statistically significant associations at $P < 0.05$ are bolded to enhance visual interpretation. \*Data are suppressed due to the limited number of outcome events.

### Secondary objective – fracture region effects

There was no evidence of effect modification between any of the anemia cohort variables (present/not present, count of co-occurring anemic conditions, clinically distinct types of anemia) and fracture region among those that sustained a fracture at the tibia (*P* for interaction, ranged from 0.189 to 0.528), suggesting that the association between fracture region and the rate of nonunion was similar between the cohorts with and without anemia, regardless of how anemia was examined. There was no evidence that anemia modified the association between tibial fracture region and nonunion risk, indicating that the regional pattern was similar in patients with and without anemia. Fracture region was therefore evaluated as the primary exposure in the combined tibial fracture cohort, with the proximal tibia as the reference. The unadjusted hazard ratio of nonunion up to 18 months after fracture was 3.18 (95 percent confidence interval, 2.67 to 3.79) for tibial shaft fractures, 0.96 (95 percent confidence interval, 0.73 to 1.26) for distal tibial fractures, and 0.72 (95 percent confidence interval, 0.38 to 1.35) for fractures at an unspecified site. After adjustment for covariates, including the presence or absence of anemia, the corresponding hazard ratios were 2.77 (95 percent confidence interval, 2.32 to 3.32), 0.93 (95 percent confidence interval, 0.70 to 1.22), and 0.78 (95 percent confidence interval, 0.41 to 1.47). Thus, the elevated risk observed for tibial shaft fractures reflects a general region-specific risk rather than a disproportionate effect of anemia at that site.

There was strong evidence that fracture region modified the association between anemia and nonunion among individuals with femoral fractures (all interaction P values <0.001). Compared with individuals without anemia, the adjusted hazard of nonunion associated with anemia was generally greatest for distal femoral fractures, followed by shaft and proximal femoral fractures, although the relative ordering of fractures at unspecified sites varied across anemia groups (**Table 5**). The regional pattern also differed according to anemia burden: hazard ratios increased with the number of anemia diagnoses at the proximal femur, whereas no corresponding increase was observed at the shaft or distal femur.

**Table 5.** Secondary objective – Adjusted^#^ hazard ratio (HR) of nonunion up to 18-months following a femur fracture for the cohorts with vs. without anemia and by anemia sub-groups based on evidence of effect modification by fracture region.

|  | Fracture region |  |  |  |
| --- | --- | --- | --- | --- |
|  | Proximal | Shaft | Distal | Site not specified |
|  | HR (95% CI) | HR (95% CI) | HR (95% CI) | HR (95% CI) |
| With any anemia | **1.20 (1.08, 1.33)** | **1.38 (1.10, 1.73)** | **2.72 (2.15, 3.46)** | 1.45 (0.98, 2.15) |
| Number of co-occurring anemic conditions |  |  |  |  |
| 1 | **1.15 (1.02, 1.29)** | **1.43 (1.12, 1.83)** | **2.87 (2.22, 3.71)** | **1.64 (1.07, 2.52)** |
| 2 | **1.21 (1.03, 1.43)** | 1.31 (0.92, 1.87) | **2.79 (1.96, 3.97)** | 1.04 (0.51, 2.12) |
| ≥3 | **1.43 (1.18, 1.74)** | 1.28 (0.84, 1.96) | **1.99 (1.21, 3.26)** | 1.36 (0.61, 3.03) |
| Nutritional anemias | **1.28 (1.10, 1.48)** | 1.33 (0.96, 1.84) | **2.10 (1.49, 2.96)** | 1.62 (0.96, 2.75) |
| Hemolytic anemias | * | * | * | * |
| Aplastic anemias | * | * | * | * |
| Other anemias | **1.19 (1.07, 1.32)** | **1.38 (1.09, 1.73)** | **2.91 (2.29, 3.70)** | 1.42 (0.95, 2.13) |

CI, confidence interval. \*Adjusted for age (as continuous), sex, insurance type, U.S. region of residence, Elixhauser Comorbidity Index score, iron deficiency, fracture type of the qualifying fracture event (closed, open, pathological), and baseline fracture at any non tibia/femur site. Statistically significant associations at $P < 0.05$ are bolded to enhance visual interpretation. \*Data are suppressed due to the limited number of outcome events.

## Discussion

This retrospective cohort study is, to our knowledge, among the largest U.S. population-based analyses to demonstrate that pre-fracture anemia is independently associated with a significantly elevated risk of nonunion following tibial and femoral fractures. A dose-response relationship was observed, with nonunion risk increasing as the number of anemia diagnoses increased, suggesting that a greater anemia burden may further compromise fracture healing. The consistency of elevated risk across most anemia subgroups, particularly nutritional and aplastic/other anemias, suggests a shared mechanistic basis, rather than a single etiology. The association between anemia and nonunion was consistently stronger for tibial than femoral fractures, even after adjustment for fracture type and comorbidities. Regional analyses further revealed that within the femur, the highest relative hazard was observed at the distal site, a pattern not replicated for the tibia, suggesting that local skeletal biology, including marrow composition and its proximal-to-distal gradient, may independently modify the relationship between systemic anemia and fracture healing outcomes.

These findings extend a growing body of evidence linking preoperative and perioperative anemia to adverse orthopaedic outcomes, including postoperative complications, transfusion requirements, and mortality^(4,5)^, by demonstrating an independent association with the longer-term outcome of nonunion. This association persisted after adjustment for comorbidities, fracture characteristics, and evidence of a fracture at another anatomical site during the preceding year. Independent of anemia status, tibial fractures carried a substantially higher nonunion risk than femoral fractures, consistent with prior population-level findings^(18,19)^. The other anemias category accounted for most patients with anemia and was clinically heterogeneous, including acute posthemorrhagic anemia, anemia associated with chronic disease, and other specified or unspecified anemias. Because these diagnoses were recorded during the year preceding the qualifying tibia or femur fracture, acute posthemorrhagic anemia could not have resulted from the index fracture itself. However, administrative claims do not establish whether the underlying hemorrhagic event was related to an earlier fracture, whether the anemia persisted until the qualifying fracture, or whether it involved iron deficiency, inflammation, or iron sequestration. The observed association in this group may therefore reflect a range of underlying conditions associated with reduced hematologic reserve or poorer overall health rather than a single biological mechanism.

Iron-deficiency anemia, the predominant diagnosis within the nutritional anemia group, was independently associated with nonunion risk, consistent with the established importance of iron for both erythropoiesis and bone metabolism. Higher dietary iron intake has been associated with preservation of bone mineral density in a dose-dependent manner^(10)^, while hemoglobin levels have been independently linked to bone mineral density in older adults^(12)^. Anemia has also been associated with increased fracture risk independent of bone mineral density^(13)^. Estimates for hemolytic and aplastic anemias were imprecise because of the small number of nonunion events in these subgroups and should therefore be interpreted cautiously. Nevertheless, skeletal complications, including reduced bone mineral density and impaired bone integrity, are well documented in sickle cell disease and related hemolytic conditions^(11)^, while intrinsic marrow failure disorders such as myelodysplastic syndromes are associated with altered bone homeostasis and skeletal abnormalities^(14,20)^.

Bone repair is metabolically demanding, relying on coordinated angiogenesis, callus maturation, and osteoblastic activity, processes critically dependent on local oxygen availability and therefore sensitive to systemic reductions in hemoglobin^(21,22)^. These processes are tightly regulated by hypoxia-responsive pathways, wherein controlled local hypoxia promotes repair while excessive or dysregulated hypoxia impairs it^(23–25)^. Systemic anemia may shift this balance beyond adaptive thresholds, exacerbating local hypoxia at the fracture site. Supporting this, we have recently demonstrated that fracture locally activates erythroid precursors in the adjacent marrow, which concentrate oxygen at the injury site through hemoglobin formation, a mechanism likely sensitive to systemic anemia^(24)^. In addition, many anemic states drive a sustained inflammatory milieu that independently disrupts the sequential phases of fracture repair^(26,27)^.

Subtype-specific mechanisms further elaborate this relationship. Iron deficiency, the predominant contributor within the nutritional anemia group, suppresses osteoblast activity while upregulating osteoclast-mediated resorption, creating a net catabolic environment unfavorable to fracture repair^(28–31)^, with additional roles in osteoblast mitochondrial function, collagen synthesis, and bone remodeling. In anemia of chronic disease and after trauma, functional iron deficiency through sequestration, rather than absolute depletion, is a predominant mechanism, present in up to 80% of operative fracture patients yet frequently undocumented^(6,8)^. In contrast, iron excess in chronically transfused conditions such as thalassemia is equally harmful, driving oxidative damage to osteoblasts and activating osteoclasts, underscoring that skeletal health is sensitive to iron dysregulation in both directions^(32,33)^, illustrating that the relationship between iron availability and bone healing is non-linear. More broadly, other disorders of erythropoiesis, including sickle cell disease and myelodysplastic syndromes, are associated with skeletal abnormalities, impaired bone integrity, and increased fracture risk^(11,14)^.

The association between anemia and nonunion was consistently stronger for tibial than femoral fractures. The tibia’s relatively limited soft tissue coverage and vascularity may render fracture healing particularly susceptible to systemic impairments in oxygen and nutrient delivery, such that anemia exerts a disproportionate effect when local biological resources are already constrained, consistent with the established higher nonunion rates at this site^(18,19)^.

Within the femur, regional analyses revealed the highest relative hazard at the distal site, a pattern not replicated for the tibia. This heterogeneity may reflect differences in marrow composition along the proximal-to-distal gradient: the proximal femur contains a greater volume of haematopoietically active red marrow, where erythroid progenitor and osteogenic cells share a more reactive niche, whereas the distal femur is proportionally richer in yellow (fatty) marrow. Evidence supports that fractures at fatty marrow sites heal less efficiently than those at red marrow sites^(34,35)^, suggesting that distal femoral healing is more dependent on systemic hematologic support and therefore more sensitive to anemia-induced perturbation of the erythroid-osteogenic niche. This relationship is bidirectional: mesenchymal stromal cell- osteoblast lineages are known to support the hematopoietic stem cell niche in human bone marrow^(35,36)^, and following fracture, erythroid precursors activated in the adjacent marrow may in turn regulate local oxygen tension and modulate the pace of osteogenesis^(24)^. Clinical observations are consistent with this framework. Fast callus appearance in tibial fracture patients with traumatic brain injury has been associated with lower erythrocyte counts and hematocrit^(37)^, suggesting that erythrocyte abundance at the fracture site exerts time-dependent influence on bone repair. Together, these data support a model in which the local erythroid environment, shaped by both systemic anemia and regional marrow composition, plays an active and dynamic role in determining fracture healing outcomes.

### Limitations

This study benefits from a large, nationally representative cohort spanning commercial and Medicare databases, enabling site- and region-specific analyses with adjustment for potential confounders and the ability to test for and quantify effect modification. Several limitations warrant acknowledgment. Anemia was identified from administrative codes rather than laboratory values, precluding assessment of severity, chronicity, or treatment response; the predominance of acute posthemorrhagic anemia codes within the aplastic/other category further introduces potential misclassification of exposure timing, as some diagnoses may reflect prior rather than truly pre-existing hematologic compromise. Bias from unmeasured confounding is possible, such as by smoking, nutritional status, medication use, undiagnosed metabolic or endocrine conditions, surgical technique, and rehabilitation adherence^(38,39)^. Data on anemia-directed treatments (iron supplementation, transfusion, or erythropoiesis-stimulating therapies) were unavailable, limiting inference regarding the potential impact of correction. The role of chronic kidney disease, a frequent driver of anemia^(40)^, as a confounder or mediator warrants dedicated evaluation in future work. As an observational study, causal inference is not possible.

### Conclusion and Clinical Implications

Pre-fracture anemia is associated with an increased risk of nonunion following tibial and femoral fractures, with a stronger and more consistent effect observed for tibial fractures, underscoring the importance of systemic host factors in fracture healing. Unlike many established risk factors, anemia is common, measurable from routine clinical data, and potentially modifiable. While causality cannot be established from this observational study, the findings support inclusion of anemia in pre-fracture or early post-fracture risk assessment, with early identification facilitating closer monitoring, multidisciplinary evaluation, and informed decision-making, particularly for higher-risk sites such as the tibia. Integration of hematology consultation and fracture liaison services could provide a coordinated framework bridging orthopaedic care with medical optimization to reduce the burden of nonunion. Non-transfusion strategies, including iron supplementation and cell salvage, represent understudied but promising avenues for anemia management in this population^(41,42)^, and transfusion thresholds should be individualized given evidence that conservative strategies are safe in stable trauma patients, but that anemia impedes early functional recovery^(43,44)^.

Future prospective studies incorporating longitudinal hemoglobin levels, iron indices, inflammatory markers, and standardized radiographic outcomes are needed to clarify the temporal relationship between anemia and impaired union. Mechanistic work should define clinically relevant thresholds of anemia that compromise bone repair and characterize interactions with hypoxia-responsive pathways central to osteogenesis and angiogenesis, ultimately informing whether targeted anemia correction can reduce nonunion rates.

## Disclosures

### Data availability statement

Data supporting the findings of this study are available within the article and supplementary materials. The underlying claims databases are subject to licensing restrictions and are therefore not publicly available.

### Ethics approval statement

The Institutional Review Board of the University of Michigan waived ethical approval for this study because the databases were de-identified before being made available to investigators.

### Patient consent statement

Patient consent was not required because all data were de-identified prior to analysis.

## Supplementary Information

**Supplementary Figure 1.**
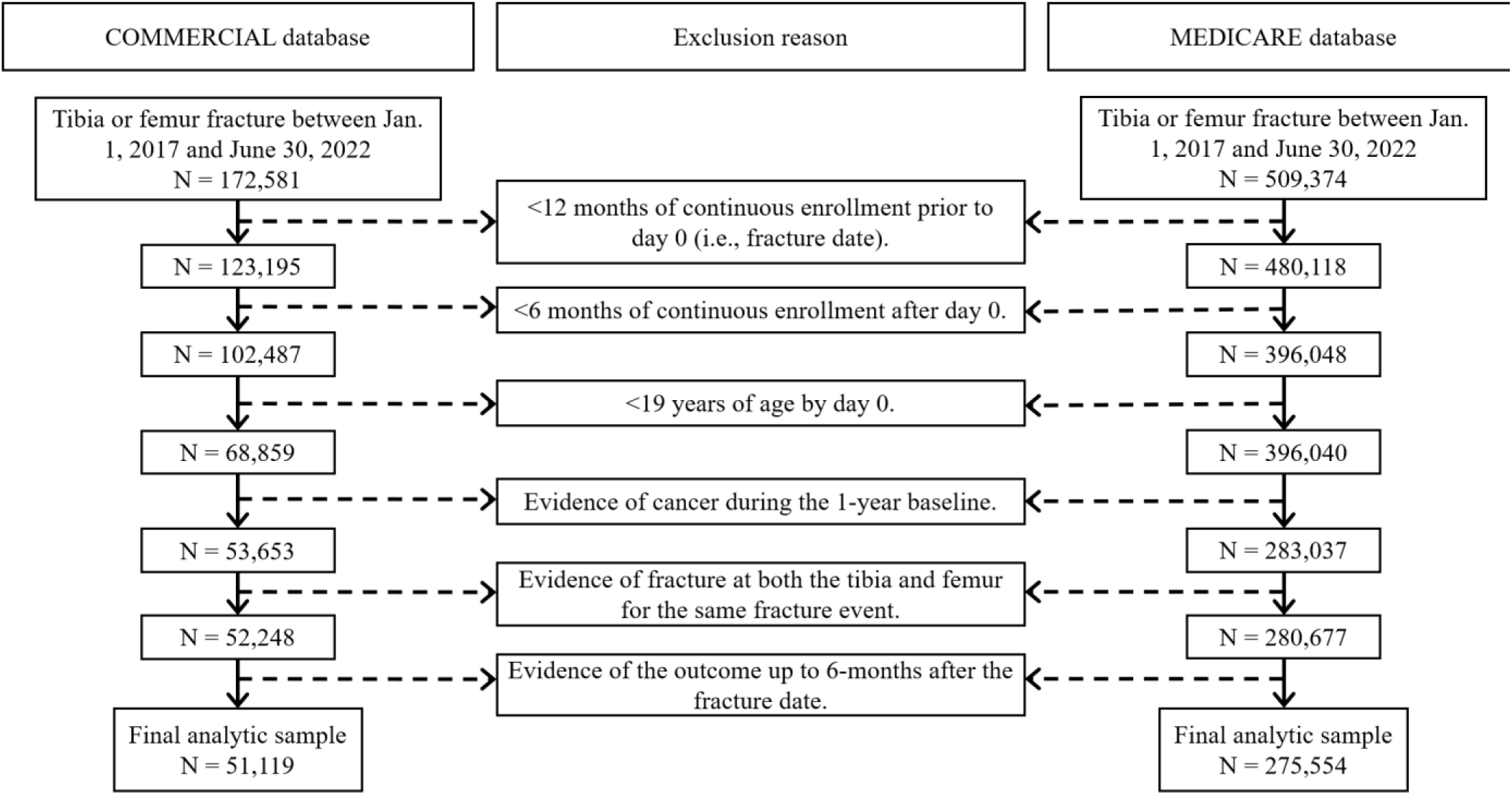
Flow chart of inclusion/exclusion criteria to derive the final analytic sample.

